# Association between Intimate Partner Violence and Nutritional Status of Children: A Systematic Review and Mata-Analysis

**DOI:** 10.1101/2021.02.19.21252077

**Authors:** Anoop Khanna, J P Singh, Neeraj Sharma

## Abstract

**Introduction:** Intimate Partner Violence (IPV) against women is a universal problem and an important social determinant of health. Studies indicate a relationship between maternal exposure to IPV and negative pregnancy and child health outcomes. The present review is aimed at doing a comprehensive review to assess the evidence for the association of IPV with the nutritional status of children.

**Methodology:** Data on the association between IPV and nutritional outcomes were extracted from 24 studies. Separate sub-group analyses were conducted for studies measuring IPV and different nutritional measures (Stunting, Underweight, and Wasting). A random-effect model was used for analysing the effect-size and the pooled effect for each subgroup.

**Findings:** The pooled estimates of ‘any violence’ [OR=1.16 (1.08-1.25)], ‘physical violence’ [OR=1.12 (1.07-1.18)] and ‘sexual violence’ [OR=1.21 (1.03-1.43)] indicated significant relationship with nutritional status of children. The relationship was found significant for stunting and underweight, but not for wasting. Values of *I*^*2*^ indicated a high level of heterogeneity across the studies on stunting and underweight.

**Conclusion:** The present review contributes to a better understanding of the nutritional outcomes for children exposed to maternal IPV. It emphasises the need to intervene for improving the well-being of these individuals as children and, subsequently, as adults.

## Introduction

Malnutrition is among the major risk factors for mortality, contributing to nearly 12 per cent of all deaths and 16 per cent of all disability-adjusted life years lost globally (1). Studies carried out in past have increased our understanding of factors contributing to child health which includes diverse public health, socio-cultural and economic factors, one of which is violence by an intimate partner (2). There is an association of undernutrition among children with marital disharmony because childcare is not only affected by a mother’s direct actions but also through her social relationships with others who facilitate the care for a child (3).

Intimate Partner Violence (IPV) against women is a universal problem, an important social determinant of health, and a major public health concern. It includes physical aggression, sexual coercion, verbal abuse and controlling behaviour. The multi-country studies indicate that nearly one-third of all women worldwide have experienced physical and/or sexual violence by an intimate partner, ranging from about 15 per cent in Japan to 71 per cent in Ethiopia (4), and such proportion was relatively high in central sub-Saharan Africa, western sub-Saharan Africa and South Asia (5). IPV is a psychosocial factor that may be associated with anaemia and underweight for women as well as their children (6). The psychological stress place by the violence on children can affect immune reactivity, predisposing children to severe and chronic infections, like diarrhea (7). Children who are exposed to maternal violence may have higher stress levels, which leads to decreased metabolic rate as well as nutritional and functional growth (8).

Literature reviews have also documented the existence of a relationship between maternal exposure to IPV and negative pregnancy and child health outcome. A systematic review and meta-analysis of IPV during pregnancy and selected birth outcomes concluded a causal relationship between IPV and low birth weight (LBW) and preterm birth (9). In another systematic review of the association between IPV and breastfeeding practices, of the 12 studies included in this review, eight found a lower breastfeeding intention, breastfeeding initiation, and exclusive breastfeeding, and a higher likelihood of early termination of exclusive breastfeeding among women exposed to domestic violence (10). Studies suggest that battered women are two to four times more likely to deliver low birth weight babies, which seriously affects the survival chances of the baby during infancy (11). An Indian study found significant differences between battered and non-battered women in terms of antenatal check-ups, safe delivery, immunization of children, birth-weight and pregnancy intention (12). A study in African context revealed that controlling behavior in marriage and experiences of all forms of IPV during a lifetime was more than four, and two times as likely to be associated with under-five mortality (13). Mothers experiencing domestic violence also tend to have poorer prenatal health, higher odds of health risk behaviors in pregnancy, less or delayed prenatal care, and higher odds of a range of poor pregnancy outcomes, including fetal death, low birth weight, and preterm birth (14). Children of well-nourished mothers (i.e., those in the normal and overweight body mass index (BMI) categories) were at lower risk of underweight, stunting, and wasting than the children of undernourished mothers.

There can be several possible explanations for higher odds of malnutrition among children of abused women. For example, domestic violence harms children’s nutrition because it is associated with a lesser ability of women to make decisions which, in turn, negatively affects the quality and quantity of the resources used towards children’s health (15). Women experiencing violence within marriage have poor status within the family that leads to weaker control over household resources, tighter time constraints, less access to information and health services, poorer mental health, and lower self-esteem. This further leads to poor health outcomes among women and children. Recent literature reviews associate direct and indirect pathways of IPV against women with child malnutrition and explore several potential mechanisms through which violence against women can affect nutritional status and other health indicators of children (16).

The literature highlights that the link between IPV and child health outcomes follows a complex pathway and needs further exploration and quantification (2). Nutritional effects of domestic violence on young children are probably high; though this relationship is less explored, and the available findings are mixed and varying (2). The purpose of the present systematic review and meta-analysis was to do a comprehensive review of the published literature to assess the evidence for the association of IPV with the nutritional status of children.

## Materials and methods

### Protocol and Registration

This systematic review is reported following the Preferred Reporting Items for Systematic Reviews and Meta-Analyses (PRISMA) guidelines and registered in the International Prospective Register of Systematic Reviews (PROSPERO - CRD42020196482). Available from: https://www.crd.york.ac.uk/PROSPERO/display_record.php?RecordID=196482

### Information Sources

The search was carried out in databases including PubMed, Scopus, Embase and Web of Science. Initial twenty pages of Google Scholar were screened for any missing article. The databases were searched from their respective start dates to March 31, 2020, using search terms tailored for each database. Bibliographic database searches and targeted hand searches were conducted to identify studies that presented a measure of association between IPV and nutritional status.

### Search strategy

To capture the breadth of study on our topic, no date limits were used in the search. Searches were conducted for English language articles only. The initial search covered the following keywords: “intimate partner violence”, “IPV”, “violence against women,” “battered women,” “domestic violence,” “domestic abuse,” “partner abuse”, “spouse abuse,” “violence,” “abuse,” “wife abuse,” “nutritional status”, “child health”, “nutritional outcome”, “stunting”, “wasting”, and “underweight”. This search strategy was modified according to the need of the specific search engines (Annexure – 1).

### Eligibility criteria

All original observational studies (cohort, case-control, and cross-sectional studies) that investigated IPV and its potential association with stunting, underweight and wasting were considered eligible. Articles that focused on special population groups, were not included in this review. Studies including physical fights, general physical assault during pregnancy, and assault without specification of the perpetrator were also excluded. Articles published only in English, in which an effect size was presented were eligible for inclusion. For studies also measuring emotional/ psychological abuse by a partner, this type of violence was included only in the systematic review but not included in the meta-analysis. After assembling the relevant studies, three categories of nutritional status of children under 5 years were established for the review: stunting, underweight and wasting.

Systematic reviews were not included in the meta-analysis as such but the original studies from these reviews were considered for inclusion. Studies that did not assess the association of IPV and nutritional status through association measures were excluded. The World Health Organization (WHO) definition of IPV - “behaviour by an intimate partner or ex-partner that causes physical, sexual or psychological harm, including physical aggression, sexual coercion, psychological abuse, and controlling behaviours”—was followed in this systematic review. However, for the meta-analyses, only studies measuring ‘any violence’ or ‘physical and/ or sexual violence’ were included.

### Data management and extraction

We imported all identified references into Zotero reference management software (17) and combined them into a single library to remove the duplicate records. A single file was imported into the Rayyan software (18) for titles and abstracts based screening as per pre-defined inclusion or exclusion criteria. A PRISMA flow diagram of the article’s selection procedure was prepared.

One reviewer (AK) reviewed the full text of all eligible articles and abstracted the relevant information to a prespecified data extraction format. Items extracted from each article were compiled into a table using Microsoft Excel. This was reviewed by the other two authors (JPS and NS) to assess and ensure the correctness of data extraction. Any inconsistencies between the reviewers were sorted out by consensus. The data extraction from eligible studies included the name of the author, method/ type of data, country, type of analysis, sample size, indicators covered (stunting, underweight, wasting), type of IPV covered (any, physical, sexual, both, emotional) and key findings. Unadjusted and adjusted odds ratios (ORs) were extracted from the selected articles. For the studies that reported raw numbers, relative risks, ORs and respective 95 per cent confidence intervals (CIs) were calculated. For the studies which did not mention the confidence interval, the same was calculated using an inverse method.

### Data Synthesis

We conducted a meta-analysis to obtain overall estimates of the effect of the intervention using a Metafor package in R software (19). We used a random-effects model. Forest plots were drawn using Log ORs and their 95 per cent CIs. Heterogeneity statistics were calculated to show how much of the total variation observed could be explained by variation between studies. ORs were considered significant when the CIs did not include 1.0, and homogeneity estimates were considered statistically significant at p<0.05. Subgroup analysis includes the analysis according to the type of violence i.e. physical, sexual, or any type of violence.

## Results

The initial literature search resulted in a total of 2335 articles. After the exclusion of the duplicates, 1041 articles were selected for the titles and abstract based screening for eligibility assessment. If insufficient data were available from the online databases to determine if a study met criteria, at this stage the article was included for full-text review. Subsequently, 142 articles were selected for full-text review. Articles whose content did not meet the criteria were eliminated. Finally, a total of 17 articles were identified for inclusion in the review. (Annexure – 2)

### Characteristics of studies

The overall characteristics of the articles included in the review were given in Table-1. A total of 17 articles were included in this systematic review. Since some of the papers analysed data of more than one country (Rico et al. (2011) studied 5 countries, Odimegwu et al. (2014) studied 3 countries and Sinha et al. (2017) studied 2 separate regions of one country) so we treated them geographically a unique study. It leads our total number of studies to 24 by 17 authors. (Table 1)

**Table 1:**
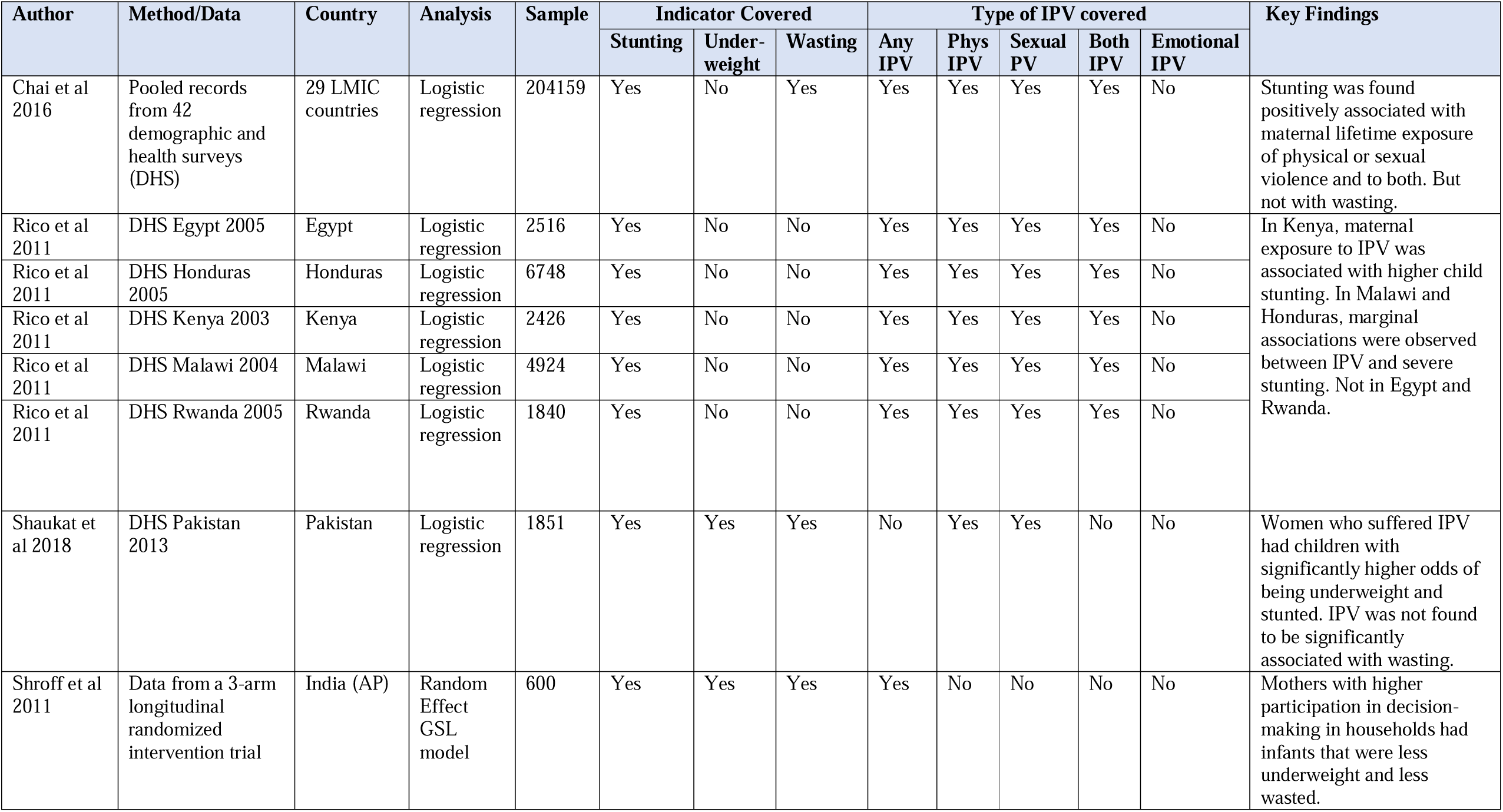
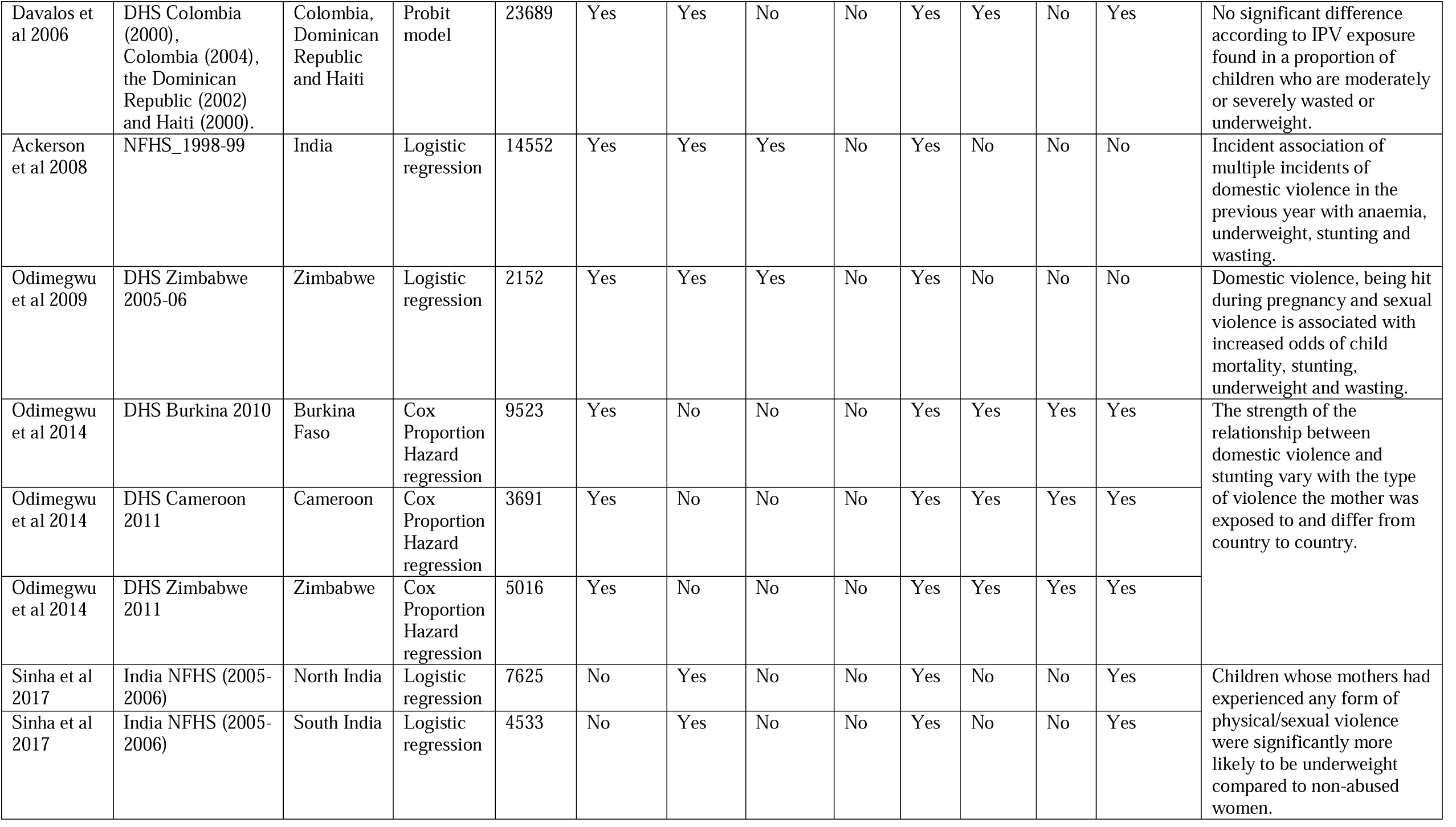
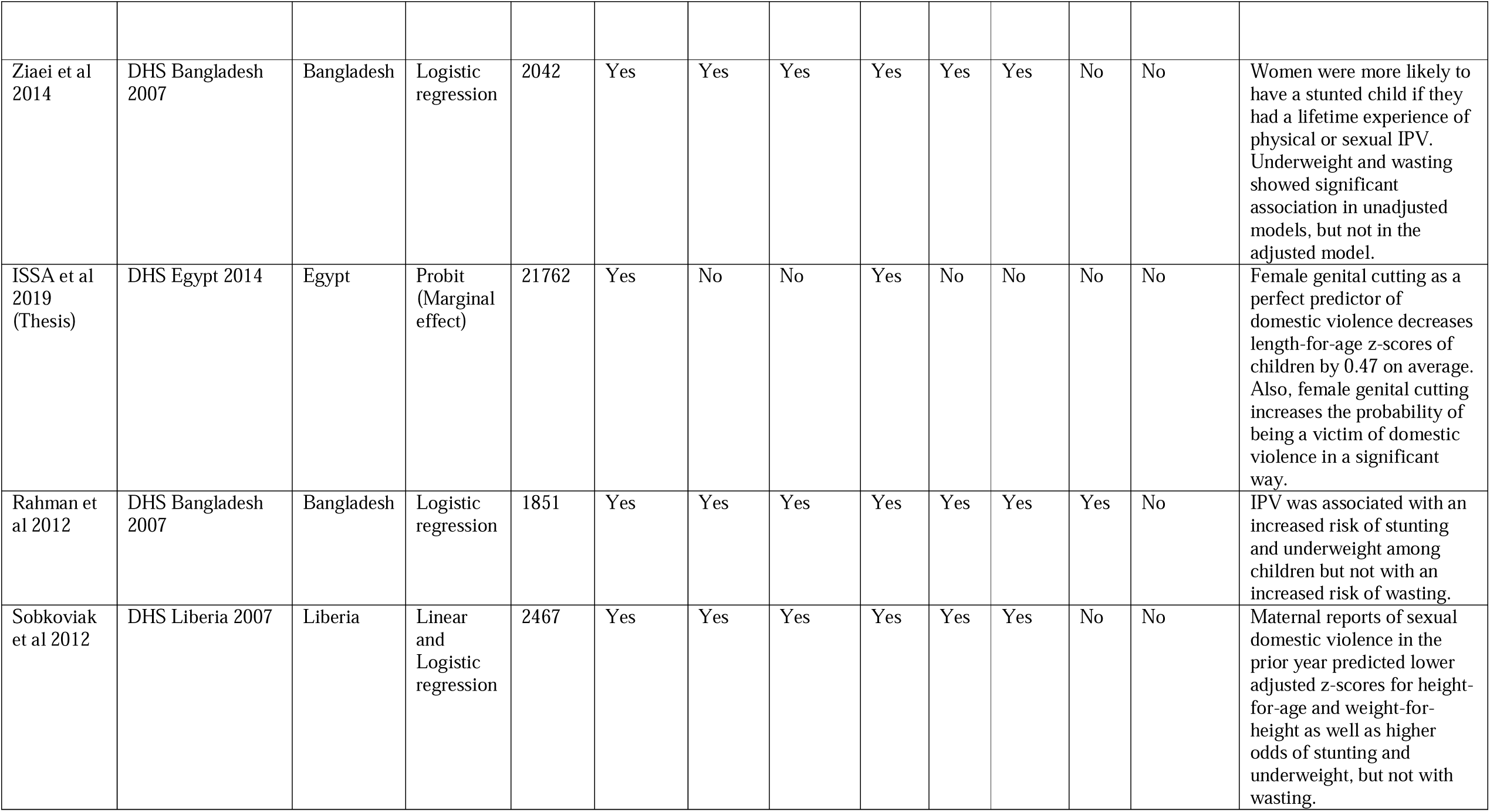
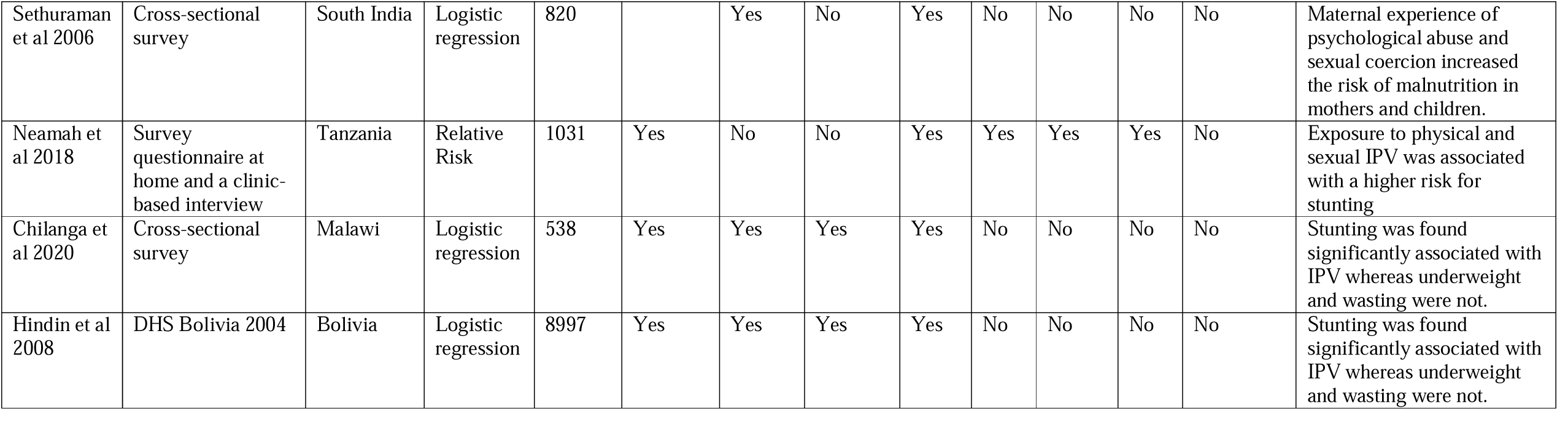
Characteristics of Studies included in the Systematic Review.

Except for one Indian study, all other studies were carried out using data from the Demographic Health Survey (DHS) conducted in different countries (20). DHS is a cross-sectional survey in which women aged 15–49 years living in a randomly selected set of households are interviewed. The studies based on the DHS mostly combined data from the domestic violence module of the DHS with data collected in the women’s questionnaire. The studies included mother-child dyads who completed the domestic violence module. One of the studies pooled data from 42 DHS in 29 Low- and Middle-Income Countries (LMICs) (21) and another one used pooled data of Colombia, the Dominican Republic and Haiti to estimate data for Latin American countries (15). Whereas one study used data from the DHS of five countries, but this study analysed each country separately rather than using pooled data of five countries (2). Similarly, one more study included three countries and presented the analysis of each country separately rather than pooling data of those countries (22). Shroff et al. used baseline data from a 3-arm longitudinal randomized education intervention trial aimed at improving the feeding, growth, and development of 3–15 months old infants (20).

Among the total 24 studies, 17 studies used logistics regression model and presented unadjusted and adjusted Odd Ratios with 95 per cent CI or standard error. Chai et al (2016) included survey fixed effects in all of the regression models to control for unobservable differences in country-specific factors as well as differences in measurement (21). To evaluate the significance of stratified associations, the authors used a pooled ordinary least squares (OLS) model with IPV covariate interaction terms. Two studies used a Probit model (marginal effect and standard error) with stunting as a dependent variable to assess the relationship with domestic violence (15)(23). These studies also used OLS (coefficient) with length-for-age z-score as a dependent variable. In one study, linear regression was used for the three continuous outcomes (z-scores for child anthropometric status), and logistic regression was used for the three binary outcomes (stunting, wasting and underweight) to estimate their relationships with the main explanatory variables (24). Shroff et al (2011) used a random effect Generalized Least Squares (GSL) model (15), while Neamah et al (2018) used relative risk as a measure of association (25).

Three important indicators of child nutrition based on anthropometric measures covered under this systematic review were: a) height for age (stunting), b) weight for age (underweight), and c) weight for height (wasting). All the studies used a similar standard definition of stunting, underweight and wasting. Out of 24 studies covered in this review, 21 studies included stunting, 13 included underweight, and 10 included wasting as the measure of nutritional status. Stunting was chosen by most of the studies because it reflects long-term nutritional deficiency and because it was the most prevalent marker of chronic malnutrition in the DHS datasets.

Regarding explanatory variables, 19 out of 24 studies included in the review captured physical violence, 15 covered sexual violence, 11 covered both physical and sexual violence as IPV variable. Only 6 studies covered emotional or psychological violence. The studies based on DHS used Conflict Tactics Scales (CTS) for measurement of IPV. Questions varied slightly by country but covered acts of physical violence ranging among most of the countries. One study measured the various dimensions of autonomy using a self-reported 47-item questionnaire, mostly on a 3-5 point Likert scale (20), the variable of experience of domestic violence was considered in the analysis. Another study used female genital cutting as an instrumental variable for domestic violence using Egyptian DHS data (23).

### Meta-analysis

A total of 19 studies were included in the Meta-analysis, for which odd ratios and confidence interval were either available or could be calculated. Four studies were excluded since they used different measures, while another one study (Chai et al 2016) was excluded because it used combined data of 29 LMICs, and some of these countries were covered by other studies included in the analysis (21).

Separate sub-group analyses were conducted for studies measuring any, physical or sexual IPV and different nutritional measure (Stunting, Underweight and Wasting). Although emotional/ psychological abuse is also an important form of IPV, inadequate standardization of its measurement makes it difficult to quantify the health effects of this type of violence. As such, it was not included in the main analysis of the present review.

Considering the high or moderate level of heterogeneity across subgroups of stunting, underweight and wasting, a random-effect model was used for analysing the effect-size and the pooled effect for each subgroup. For presenting the effect size, forest plots were created for each type of violence and their subgroups based on Log Odds Ratios and their 95 per cent confidence intervals. To depict the publication bias, funnel plots were created, while Egger’s test was applied for its quantitative assessment, which regresses the standardized effect sizes on their precisions.

Figure 1 (A,B,C) presents the forest plot indicating the effect size of each study as well as the pooled effect, and table 2 presents the summary results. The pooled estimates of OR 1.16 (1.08-1.25) indicating a significant relationship between any violence and nutritional status of children. The subgroup analysis indicated that the relationship was significant in pooled estimates of stunting and under-weight, while the relationship was not significant in case of wasting with OR 1.11 (0.96-1.28). The overall heterogeneity was found to be 70.1 per cent (*I*^*2*^) across the studies on any violence. It was specifically high regarding stunting (*I*^*2*^=86.5 per cent). The Cochran’s Q test was statistically significant at five per cent level of significance, suggesting the evidence for the heterogeneity between the included studies included for any violence. However, the studies on underweight and wasting were not heterogeneous.

**Table 2:** Summary Results of Meta-analysis.

|  | **Group/ Sub-group** | **Random effects Log OR (95% CI)** | **Random effects OR (95% CI)** | **Cochrane's Q (P-value)** | **Number of studies** | **I<sup>2</sup>-Value (%)** |
| --- | --- | --- | --- | --- | --- | --- |
| **A** | ***Any Violence*** |  |  |  |  |  |
|  | Stunting | 0.16 (0.05-0.28) | 1.18 (1.05-1.32) | 51.48 (0.00) | 9 | 86.5 |
|  | Underweight | 0.17 (0.07-0.27) | 1.19 (1.07-1.31) | 2.61 (0.46) | 3 | 0.0 |
|  | Wasting | 0.10 (-0.04-0.24) | 1.11 (0.96-1.28) | 0.75 (0.86) | 3 | 0.0 |
|  | Overall | 0.15 (0.08-0.22) | 1.16 (1.08-1.25) | 61.91 (0.00) | 17 | 70.1 |
| **B** | ***Physical Violence*** |  |  |  |  |  |
|  | Stunting | 0.14 (0.06-0.22) | 1.15 (1.06-1.24) | 40.85 (0.00) | 12 | 66.3 |
|  | Underweight | 0.10 (0.03-0.18) | 1.11 (1.03-1.20) | 27.64 (0.00) | 8 | 70.3 |
|  | Wasting | 0.06 (-0.10-0.22) | 1.06 (0.90-1.24) | 10.73 (0.06) | 5 | 60.7 |
|  | Overall | 0.11 (0.06-0.16) | 1.12 (1.07-1.18) | 83.47 (0.00) | 27 | 77.0 |
| **C** | ***Sexual Violence*** |  |  |  |  |  |
|  | Stunting | 0.26 (0.08-0.44) | 1.30 (1.08-1.55) | 116.84 (0.00) | 10 | 85.7 |
|  | Underweight | 0.32 (-0.13-0.78) | 1.38 (0.87-2.18) | 129.46 (0.00) | 3 | 96.8 |
|  | Wasting | -0.13 (-0.42-0.16) | 0.88 (0.66-1.17) | 8.34 (0.04) | 3 | 62.4 |
|  | Overall | 0.19 (0.03-0.36) | 1.21 (1.03-1.43) | 260.89 (0.00) | 18 | 97.1 |

**Table 3.** Egger’s Regression Test for Funnel Plot Asymmetry.

| **Type of Violence** | **z-test value** | **p-value** |
| --- | --- | --- |
| Nutrition – Any | 2.1935 | 0.0283 |
| Nutrition – Physical | 0.5609 | 0.5749 |
| Nutrition – Sexual | -0.4504 | 0.6525 |

**Figure 1(A):**
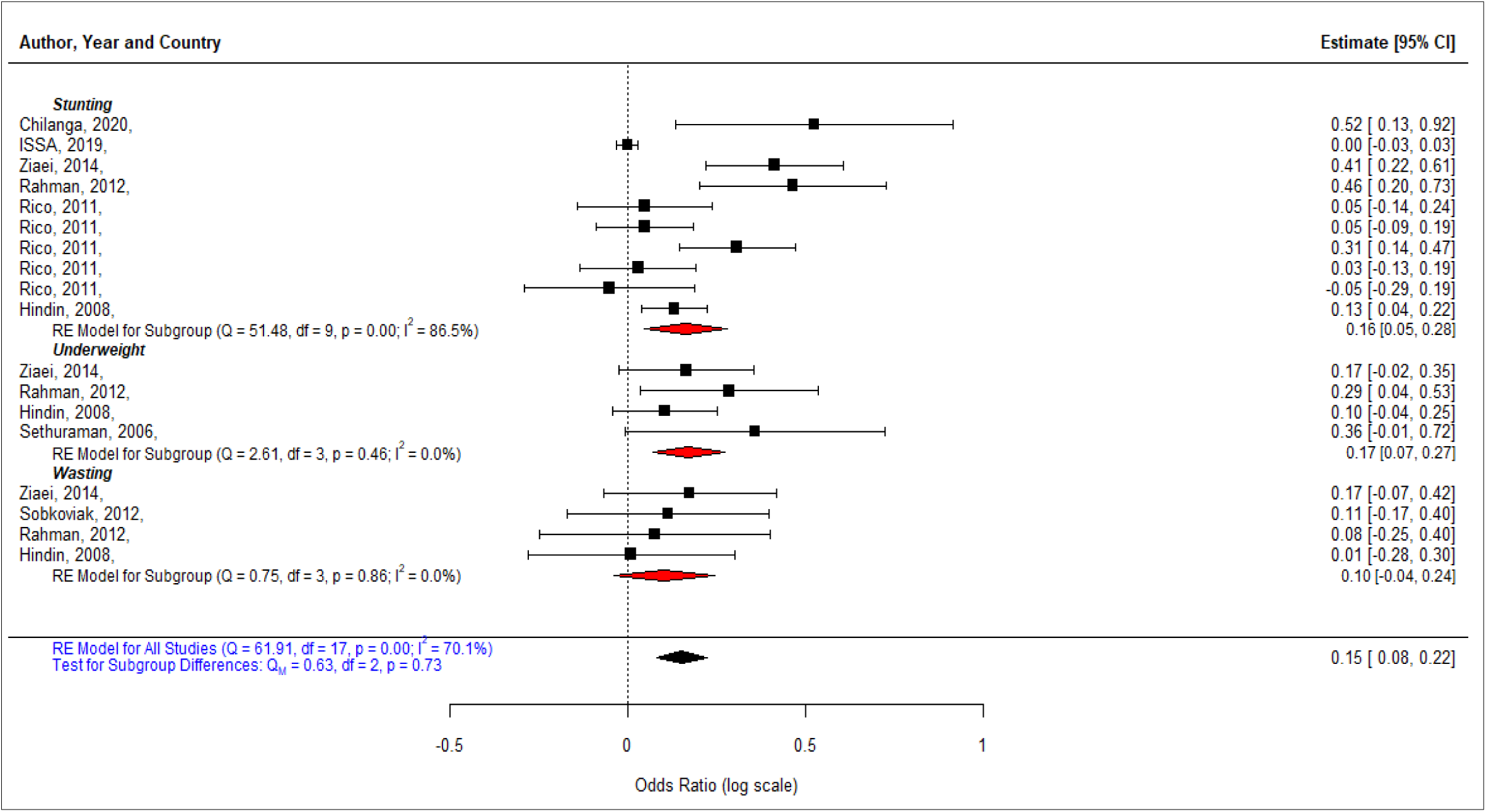
Meta-analysis results of the relationship between Any IPV and Nutritional status. Note: Meta-analysis is by random-effects, and 95% confidence intervals (CI) are shown, along with weights for each estimate. Box size is proportional to study weight, and black lines represent 95% CIs. Summary estimates for each panel are shown as diamonds.

**Figure 1 (B):**
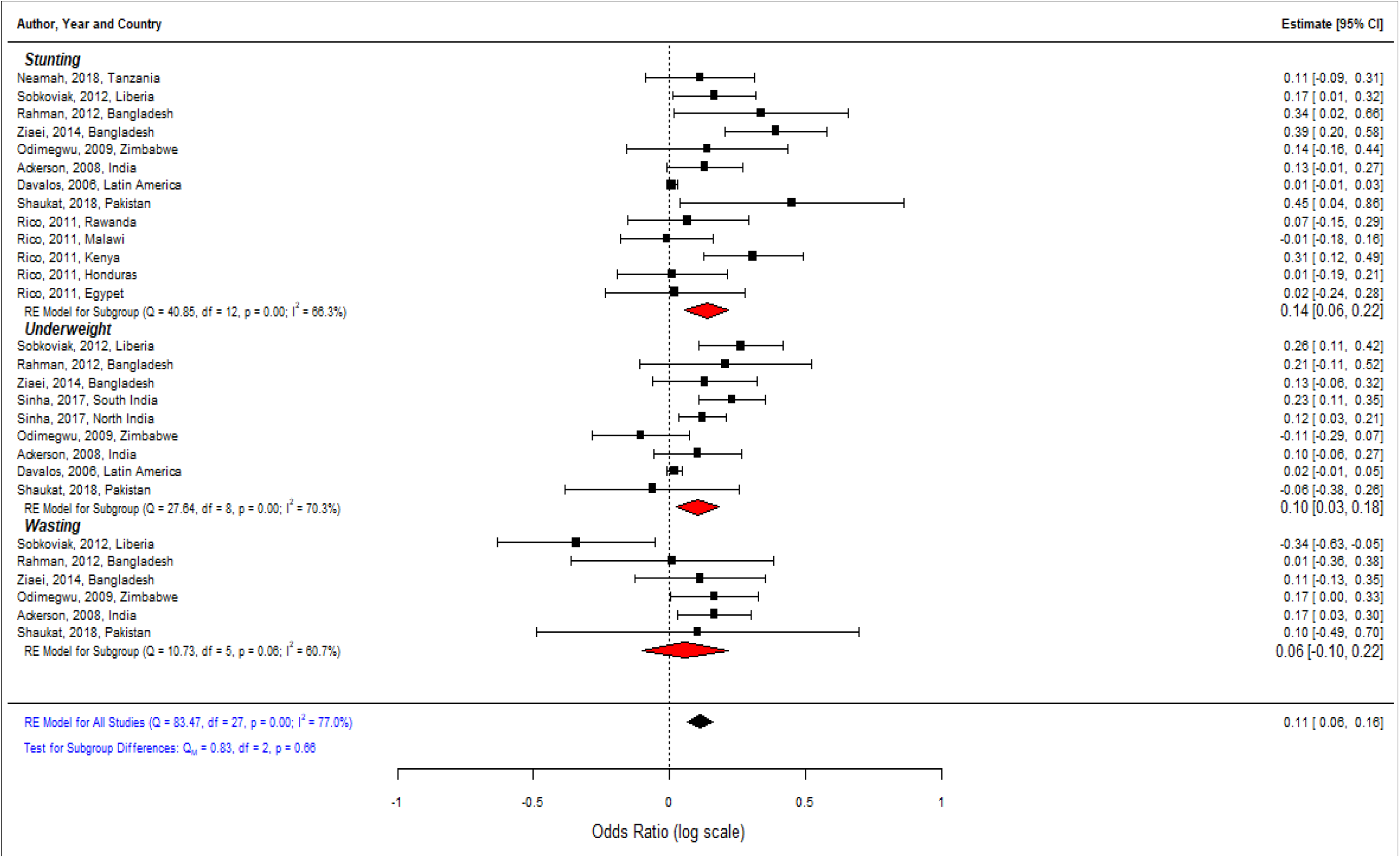
Meta-analysis results of the relationship between Physical IPV and Nutritional status. Note: Meta-analysis is by random-effects, and 95% confidence intervals (CI) are shown, along with weights for each estimate. Box size is proportional to study weight, and black lines represent 95% CIs. Summary estimates for each panel are shown as diamonds.

**Figure 1 (C):**
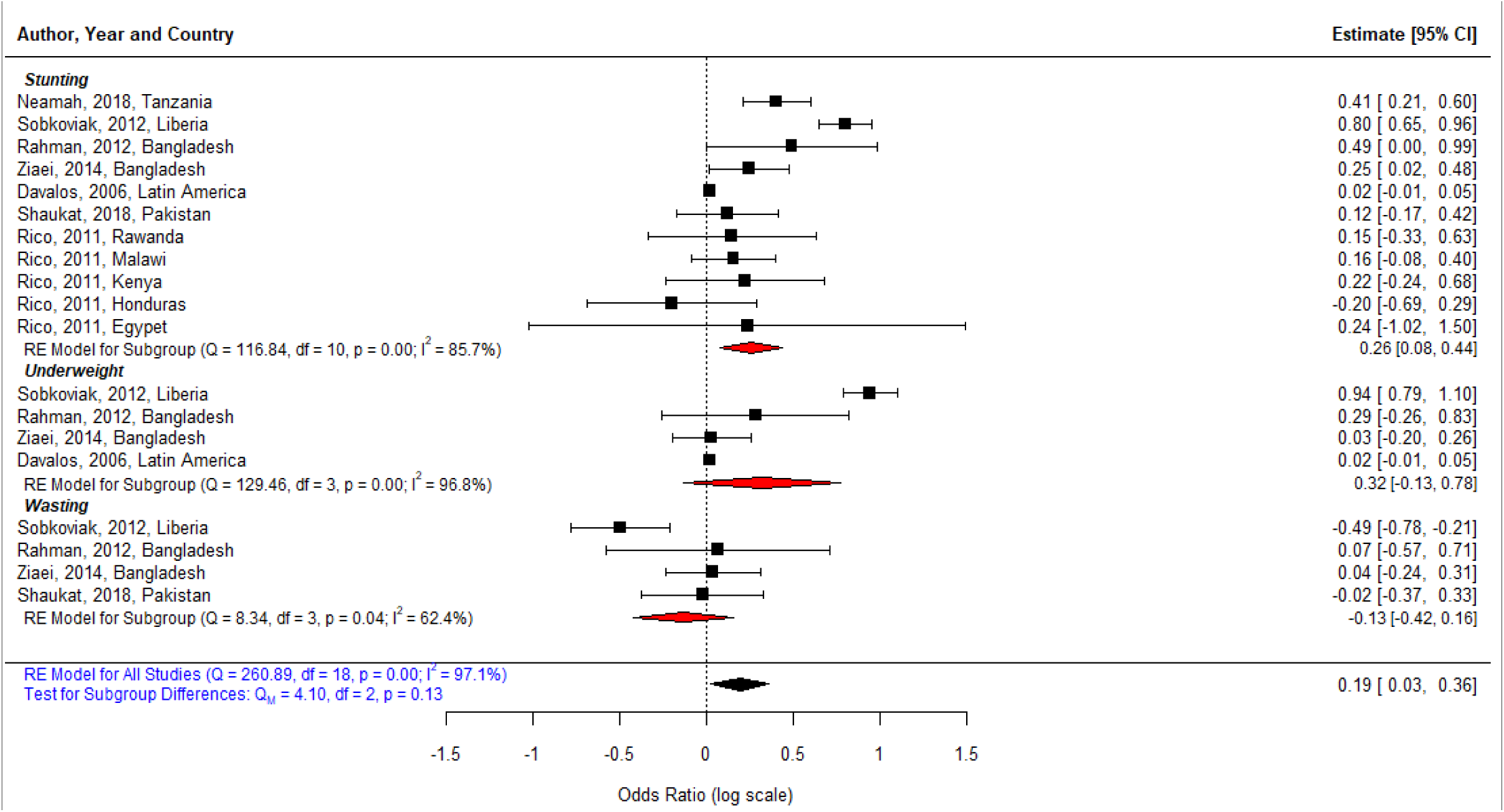
Meta-analysis results of the relationship between Sexual IPV and Nutritional status. Note: Meta-analysis is by random-effects, and 95% confidence intervals (CI) are shown, along with weights for each estimate. Box size is proportional to study weight, and black lines represent 95% CIs. Summary estimates for each panel are shown as diamonds.

The studies on physical violence also indicated a significant relationship between the experiences of physical violence and nutritional status with OR 1.12 (1.07-1.18). The relationship was significant regarding the studies on stunting and underweight, while not regarding the studies on wasting with OR 1.06 (0.90-1.24). Overall, there was a high level of heterogeneity (*I*^*2*^=77 per cent) among the studies covering physical violence, though the subgroup analysis indicated moderate (between 60.7 to 70.3 per cent) level of heterogeneity among the subgroups of stunting, underweight and wasting.

A similar pattern was found regarding sexual violence, as the pooled estimates of effect size indicated a significant relationship between experiences of sexual violence among women and nutritional status of their children in terms of stunting and underweight. Overall, the studies related to sexual violence had a very high heterogeneity (*I*^*2*^=97.1 per cent). The level of heterogeneity was found to be high among the subgroups of stunting and underweight, while it was moderate in case of wasting. The Cochran’s Q test was found to be significant (p<0.05) for all subgroups confirming the heterogeneity among the studies across all subgroups.

### Heterogeneity and Publication Bias

The funnel plot (Figure 2 - A, B and C) with pseudo 95 per cent confidence interval was plotted to assess the publication bias in the studies included in the meta-analysis, separately for any violence, physical violence and sexual violence. The plot of any violence indicated some publication bias, which is mainly due to two small sample studies (26,27). However, the funnel plots of physical violence and sexual violence did not indicate publication bias. Egger’s regression test (Table 2) also suggests that there was no conclusive evidence of publication bias, particularly regarding physical violence and sexual violence.

**Figure 2(A):**
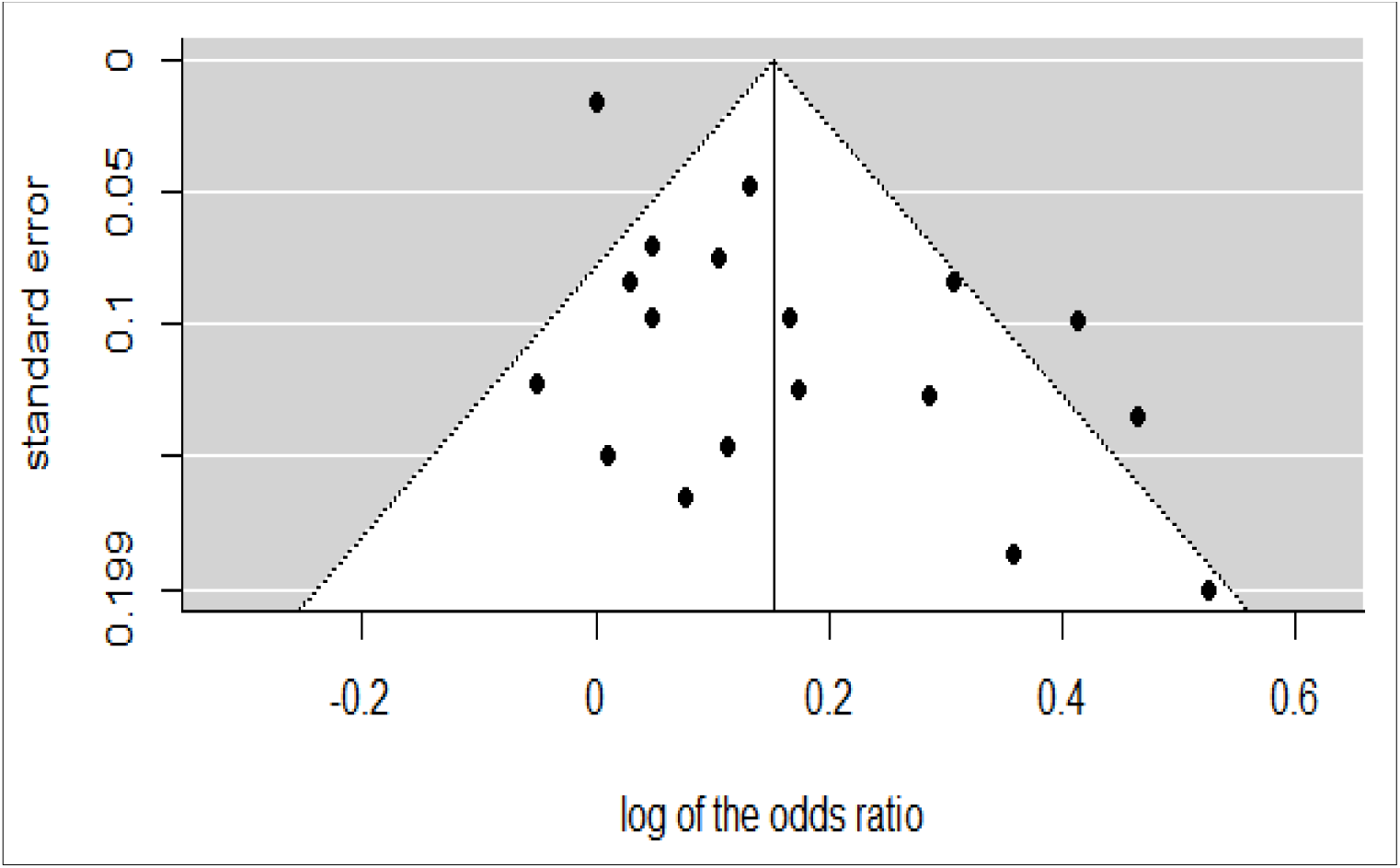
Funnel Plot: Any IPV.

**Figure 2 (B):**
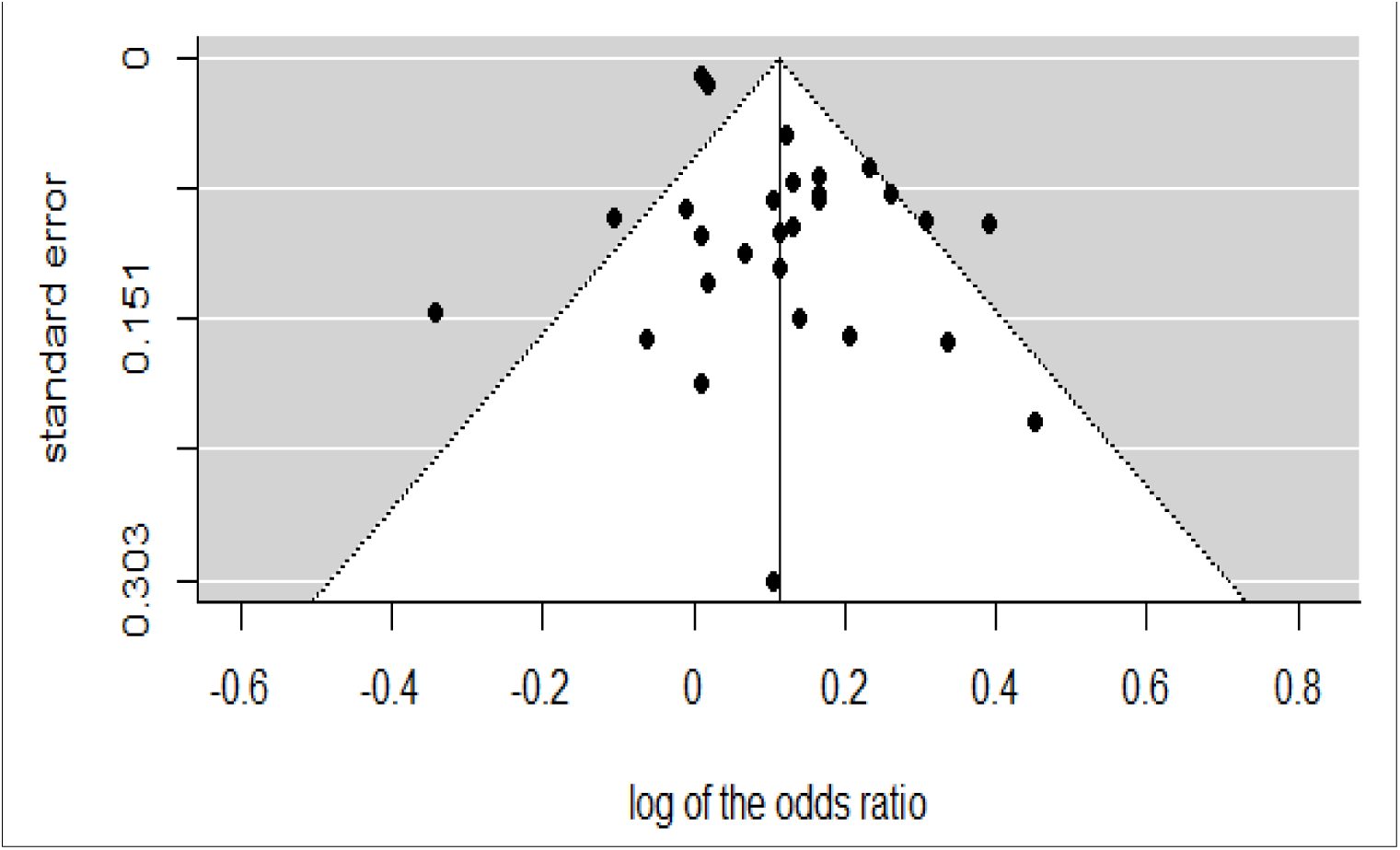
Funnel Plot: Physical IPV.

**Figure 2 (C):**
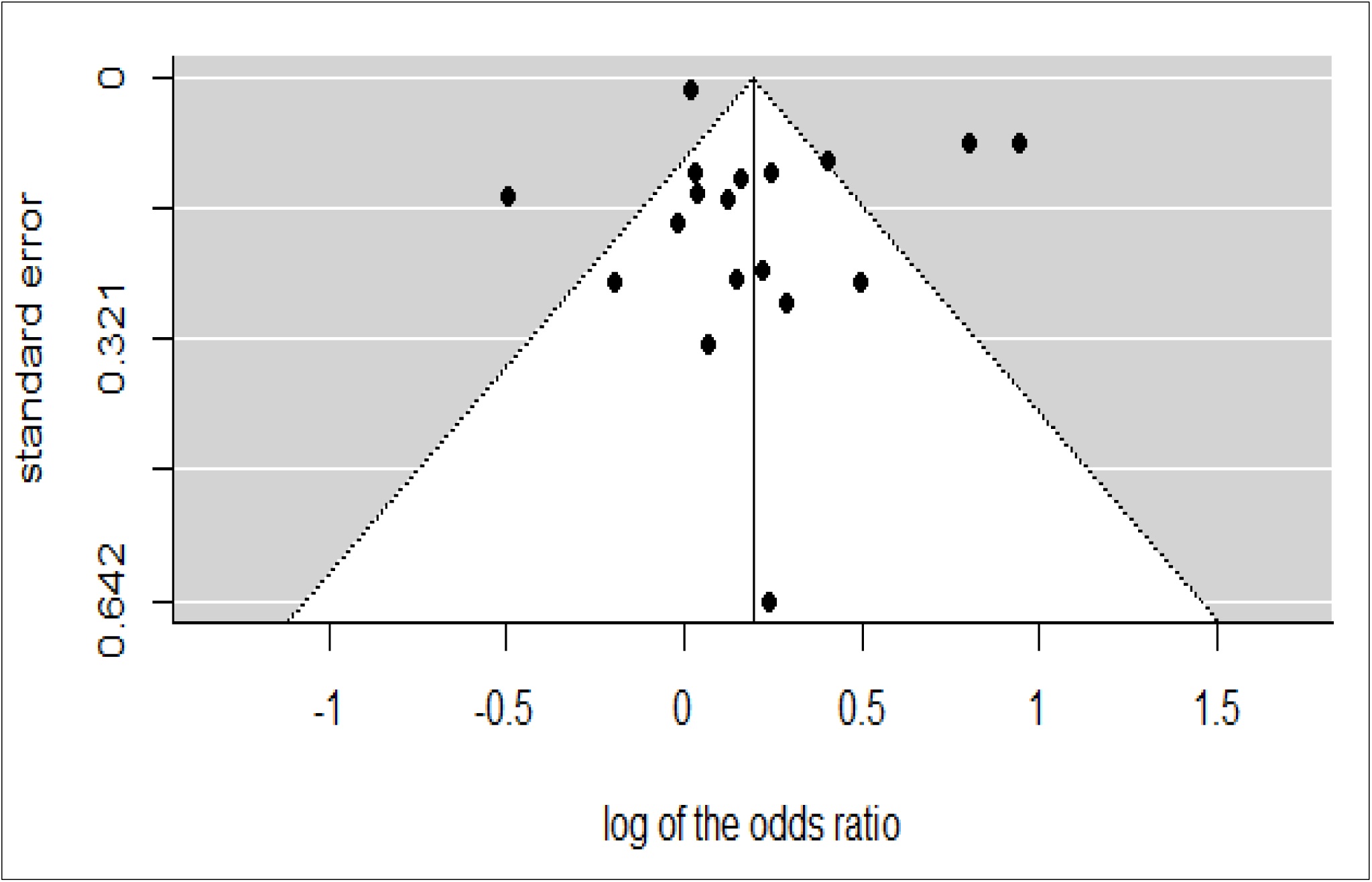
Funnel Plot: Sexual IPV.

## Discussion & Conclusion

The individual studies included in this analysis, as well as the pooled effect size of different subgroups, indicated a significant relationship between the experience of women about any, physical or sexual IPV and the nutritional outcome in terms of stunting and underweight. The study provides evidence that though the nature of IPV depends on the social, demographic and cultural context in which it occurs, there is no reason to assume that its association with children’s health and well-being is restricted to few settings only. While the rates and forms of violence may differ across the nations, their consequences on child health outcomes are similar (4,5).

An important finding is that the any, physical and sexual violence were significantly related to the stunting and underweight, but none of these indicated relationships with the wasting. Some studies found a small negative relationship of IPV with wasting, though the authors concluded that the result may be related to survivor bias in the context of a cross-sectional analysis (21). Moreover, fewer studies have tried to assess the relationship of IPV with wasting as compared to stunting and underweight.

Although all the studies have made adjustments for covariates and confounding, these variables differ across the countries. Most of the studies included the socioeconomic and demographic characteristics as covariates, which include maternal age, employment status, level of education, marital status, partner’s level of education, rural/ urban residence, wealth quintile, number of children younger than five years in the household and the child’s age. In a study, maternal height was retained in all statistical models as it is a strong predictor of child height (28).

The systematic review indicated that although most of the studies have captured physical and sexual violence, very few studies have included emotional or psychological violence to examine its association with nutritional outcomes. Several studies have provided evidence about the linkage between maternal depression and nutritional outcomes of children. In a meta-analysis, authors presented that children whose mothers had depression were 1.4 times more likely to be stunted than the children of mothers who were not depressed (29). Emotional and psychological IPV can also cause maternal depression affecting a woman’s ability to care for her child, which may further contribute to childhood malnutrition (29).

The persisting inequality between man and woman and the lower status of women is the major driving force behind the nutritional status and health of children. Initiatives to advance women’s autonomy, through access to education and economic opportunities may offset the risk of intimate partner violence (21). One study found that a lack of maternal autonomy was significantly associated with an increased likelihood of child stunting (30). There is a number of studies that found positive associations of increases in women’s healthcare autonomy using specifically child’s height-for-age scores as outcome measures (31,32). Promoting joint decision-making (of partners) could be an important public health intervention to empower women while avoiding domestic violence (33). Further longitudinal research is needed to understand the effect of autonomy over the life course on the children’s nutritional status.

Our literature review strongly suggests that reducing the maternal experience of spousal violence is an important goal for improving a child’s health and survival. The review emphasizes the need for a better understanding of the nutritional outcomes for children exposed to IPV, which in turn will require a clearer conceptualization of the causal pathway linking the two. Longitudinal studies are needed to investigate the influence of potential mechanisms mediating the association between IPV and child nutritional outcomes. Such knowledge ultimately will enhance our ability to design interventions to improve the well-being of these individuals as children and, subsequently, as adults.

The present findings contribute to the growing body of evidence showing that IPV can also compromise children’s growth, supporting the need to incorporate measures to address IPV in child health and nutrition programmes and policies. It shows the importance of collating and analysing systematically the information available on the less explored but critical issues like the association between IPV and nutrition levels.

### Limitations

A few limitations should be considered while interpreting the findings from this review and analysis. First is the exclusion of grey literature, such as reports and conference abstracts, which may have introduced an element of publication bias. Most of the studies used DHS data for analysis, and all were based on cross-sectional data. In cross-sectional studies, it is difficult to establish a cause and effect relationship. It is possible that intimate partner violence was the result of child growth or malnutrition, though such reverse causality seems relatively unlikely (21). Further analysis, with longitudinal data, may provide insights on the temporal effects of exposure to such violence on our outcomes of interest.

Another disadvantage with the cross-sectional design is the potential for survivor bias, which may have resulted in the apparent increase in the risk of wasting among children whose mothers had experienced intimate partner violence (21). There could be under-reporting of domestic violence due to social stigma, privacy concerns, and sensitive nature of information (34). Recall bias might have been introduced because women were asked to provide retrospective information regarding their experience of domestic violence. Further, although the studies have applied a rigorous adjustment for confounding factors, and all have presented adjusted Odd Ratios, there might be residual confounding. Despite these shortcomings, we believe this review represents a meaningful contribution to the existing knowledge about the association between IPV and nutritional outcomes.

## Supporting information

Annexure 1_Search Strategy

Annexure 2_PRISMA flow diagram

## Data Availability

All data generated or analyzed during this study were included in this published article [and its Additional files].

## Declarations

### Ethics approval and consent to participate

This systematic review does not require ethical clearance as it used secondary data which is already in the public domain. Also, this study has not used any unit-level data.

### Consent for publication

Not applicable

### Author’s Contribution

AK and JPS contributed in the designing, data extraction and data synthesis. NS analysed the data and AK, JPS, NS interpretation and drafted the manuscript. All authors have reviewed the manuscript critically for important intellectual content and have given final approval of the version to be published.

