## Supplementary material for "Association between Intimate Partner Violence and Nutritional Status of Children: A Systematic Review and Mata-Analysis": Annexure 1_Search Strategy

A) **PubMed**

(((women[tiab] OR “against women”[tiab] OR “battered women”[tiab] OR domestic[tiab] OR spouse[tiab] OR partner[tiab] OR wife[tiab] OR “Intimate Partner”[tiab]) AND (violence[tiab] OR abuse[tiab]) OR IPV[tiab]) AND (((malnutrition[tiab] OR nutrition*[tiab] OR Undernutrition[tiab]) AND (status[tiab] OR outcome[tiab])) OR “child health”[tiab] OR stunting[tiab] OR wasting[tiab] OR underweight[tiab]))

B) **SCOPUS**

TITLE-ABS ( ( ( women OR "against women" OR "battered women" OR domestic OR spouse OR partner OR wife OR "Intimate Partner" ) AND ( violence OR abuse ) OR ipv ) AND ( ( ( malnutrition OR nutrition* OR undernutrition ) AND ( status OR outcome ) ) OR "child health" OR stunting OR wasting OR underweight ) )

C) **Embase**

((women:ab,ti OR 'against women':ab,ti OR 'battered women':ab,ti OR domestic:ab,ti OR spouse:ab,ti OR partner:ab,ti OR wife:ab,ti OR 'intimate partner':ab,ti) AND (violence:ab,ti OR abuse:ab,ti) OR ipv:ab,ti) AND ((malnutrition:ab,ti OR nutrition*:ab,ti OR undernutrition:ab,ti) AND (status:ab,ti OR outcome:ab,ti) OR 'child health':ab,ti OR stunting:ab,ti OR wasting:ab,ti OR underweight:ab,ti)

D) **Web of Science**

AB=(((women OR “against women” OR “battered women” OR domestic OR spouse OR partner OR wife OR “Intimate Partner”) AND (violence OR abuse) OR IPV) AND (((malnutrition OR nutrition* OR Undernutrition) AND (status OR outcome)) OR “child health” OR stunting OR wasting OR underweight))
