## Supplementary material for "Association between Intimate Partner Violence and Nutritional Status of Children: A Systematic Review and Mata-Analysis": Annexure 2_PRISMA flow diagram

**Annexure – 2: Flowchart of studies included in the review (Based on PRISMA[1] framework).**

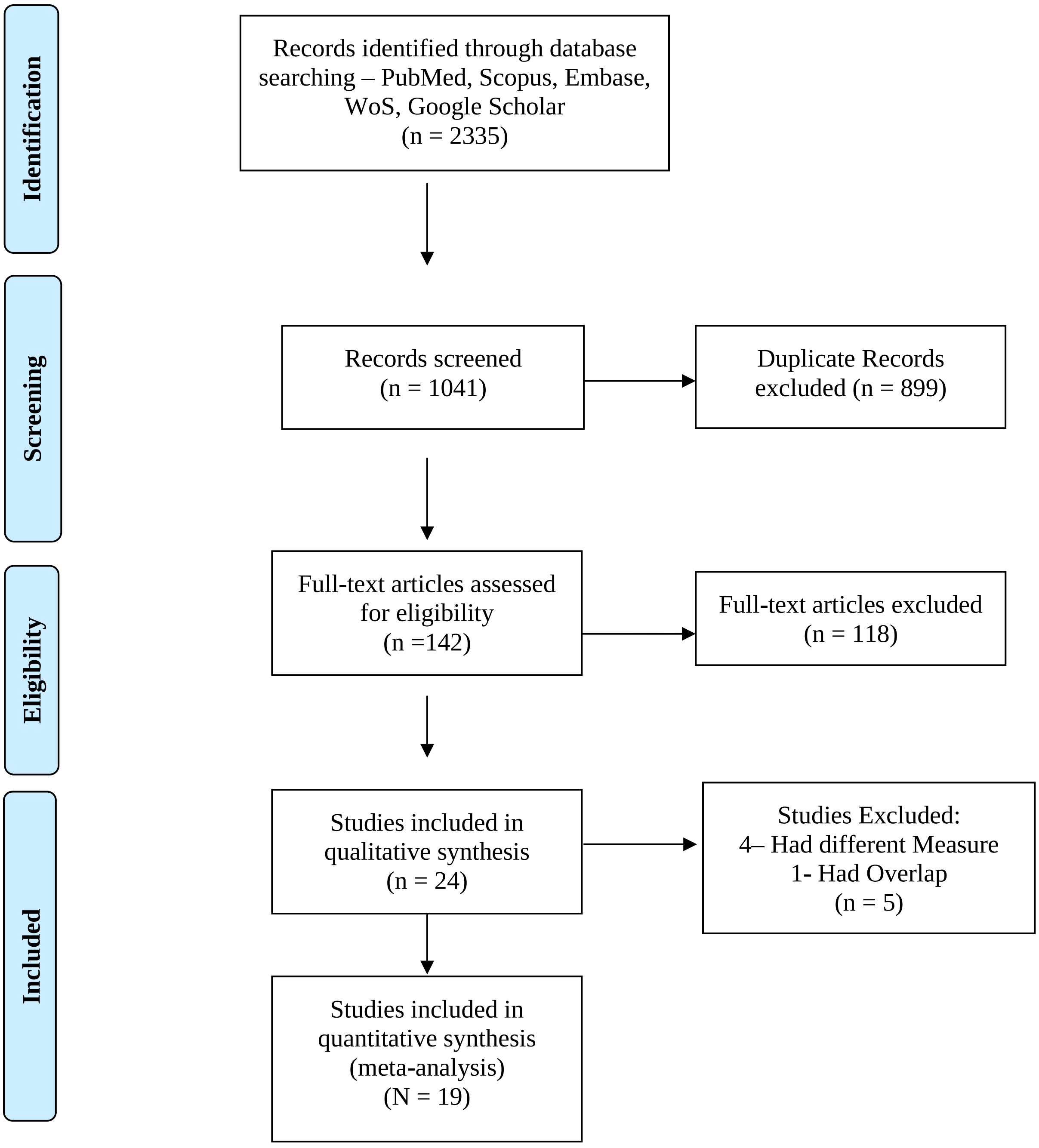


[1] **Note:** Adapted from: Moher D, Liberati A, Tetzlaff J, Altman DG, The PRISMA Group (2009). Preferred Reporting Items for Systematic Reviews and Meta-Analyses: The PRISMA Statement. PLoS Med 6(7): e1000097. doi:10.1371/journal.pmed1000097
